# Minimal measurement strategies for cardiometabolic risk classification within the Positive Health framework: A cross-sectional analysis of NHANES 1999–2004 data

**DOI:** 10.64898/2026.08.07.26359959

**Authors:** Kerstin A. Schorr, Tim J. van den Broek, Meike van den Eijnden, Femke P.M. Hoevenaars, Suzan Wopereis

## Abstract

**Background:** Large-scale prevention and population health monitoring require measurement approaches that are both feasible and informative. Although several self-measurable anthropometric and fitness indicators have been associated with cardiometabolic risk, it remains unclear whether combining multiple measurements provides meaningful improvements over simpler approaches. We evaluated whether a parsimonious set of self-measurable indicators can achieve classification performance comparable to a full candidate set and quantified the incremental value of additional measurements.

**Methods:** Using data from 8,275 adults in the NHANES 1999–2004 cohorts, we evaluated a predefined minimal set of four self measurable anthropometric and fitness indicators (body mass index (BMI), waist to height ratio (WHtR), mid upper arm circumference (MUAC), and VO2max (as a proxy for the 6 minute walk test) as candidate indicators of cardiometabolic risk. Their ability to reflect underlying clinical risk factors related to adiposity, glucose and lipid metabolism, and physical fitness was assessed using nested logistic regression models, likelihood ratio tests, discrimination metrics, and decision tree analyses.

**Results:** WHtR consistently showed the strongest discriminative performance, with ΔPR-AUC values for BMI versus WHtR ranging from -0.002 to -0.037, and emerged as the primary splitting variable. Adding BMI to WHtR resulted in small gains in PR-AUC for most outcomes, ranging from 0.000 to 0.008, except for triglycerides where the gain was larger (ΔPR-AUC=0.039). Further inclusion of MUAC and VO2max provided limited additional value overall, with evidence of variation across outcomes and sex stratified analyses.

**Conclusion:** Most classification performance was achieved using a limited number of simple self-measurable indicators, with little additional benefit from incorporating further measurements. These findings suggest that parsimonious measurement strategies may provide a feasible approach for cardiometabolic risk classification in population health and prevention settings while reducing measurement burden.

**Key Messages:** *What is already known on this topic:*

- Multiple anthropometric indicators have been associated with cardiometabolic risk
- It remains unclear which indicator or which combination of indicators provide meaningful improvements in risk classification

*What this study adds:*

- In this study we systematically evaluate the incremental value of BMI, WHtR, MUAC and VO2max to quantify their incremental contribution to classification of multiple cardiometabolic outcomes
- WHtR consistently showed the strongest classification performance, while additional measurements provided limited benefit

*How this study might affect research, practice or policy:*

- This study suggests that a parsimonious measurement strategy may be sufficient for classification of cardiometabolic risk
- Simpler, resource-efficient measurement strategies could alleviate measurement burden while maintaining risk classification

## 1. Introduction

Noncommunicable diseases (NCDs), such as cardiovascular diseases, type 2 diabetes, and cancer, remain a leading cause of mortality worldwide. These diseases continue to place a significant burden on healthcare systems. Key risk factors that are related to NCDs include adiposity, metabolic dysfunction and reduced physical fitness. Early recognition and assessment of these risk factors are essential for effective prevention of NCDs.

In recent years, healthcare and policy initiatives have increasingly emphasized a shift from curative towards preventive care, aiming to reduce the burden of NCDs on healthcare costs^1,2^. Health is increasingly recognized as a multidimensional concept, evolving beyond the mere absence of disease to encompass physical, social, and emotional well-being^3–5^. This definition consists of two equally important components: a psychosocial and contextual component and a physiological component, which addresses objective and functional measurements of health^5–7^. Building on this broader perspective, the concept of positive health (PH) was introduced by Huber et al. in 2016, defined as “the ability to adapt and to self-manage in the face of social, physical, and emotional challenges”^8^. This definition, developed through input from patients, healthcare providers, citizens, and researchers, includes six dimensions: physical function, mental function, spiritual/existential dimension, quality of life, daily functioning, and social participation^4^.

To operationalize this concept, dialogue tools such as My Positive Health (MPH) have been adopted in the Netherlands to assist practitioners in conversations about PH. However, there is an increasing need to also quantify and evaluate the impact of health-promoting interventions on aspects of PH^3^. In response, previous research has developed instruments to quantify PH, that may provide researchers, healthcare professionals, insurers, and policymakers with valid information on the effectiveness of PH and related interventions^7^. A context-sensitive positive health questionnaire has evolved from the MPH dialogue tool, including eight factors: mental relaxation, social acceptance, wellbeing, vitality, social support, financial resources, health literacy and mobility^9^. Despite its great potential for measurement of context-sensitive health, this instrument currently lacks measurements for the physiological domain of health, in particular objective indicators for metabolic and cardiovascular health.

In literature, various anthropometric measurements have been suggested being indicative of distinct aspects of physiological health. However, a systematic evaluation of which measurements contribute meaningfully to risk classification and therefore could be scalable in the public health context is currently still missing^10–13^.

To address this gap, we aimed to evaluate a predefined *minimal* set of simple, valid, at-home measurements that capture key aspects of physiological health and complement existing Positive Health (PH) instruments. Based on expert consultation and established health standards, we selected body mass index (BMI), waist-to-height ratio (WHtR), mid-upper arm circumference (MUAC), and a proxy of cardiorespiratory fitness (6-minute walking test) as candidate indicators. Using data from the National Health and Nutrition Examination Survey (NHANES), we evaluated the extent to which these indicators reflect underlying clinical cardiometabolic risk and assessed their incremental contribution when combined. Our goal was to evaluate the extent to which a reduced set of practical, self-measurable indicators can achieve classification performance comparable to the full candidate set, and to quantify the incremental value of adding measurements, while maintaining feasibility for self-assessment within the Positive Health framework.

## 2. Methods

This study is a predictive modelling study aiming to evaluate classification performance and variable prioritization. To evaluate the contribution of a predefined minimum set of simple, valid, at home anthropometric indicators to cardiometabolic risk, we first selected four self-measurable measurements based on existing health standards and expert input. We then linked these indicators to clinical risk factors using NHANES data and assessed the relative and incremental contribution of each measurement to cardiometabolic risk.

### 2.1. Definition of a minimal set of simple, valid, self-measured, at-home indicators for physiological health

The goal was to define a candidate set of valid, objective, and scientifically supported anthropometric measurements that could feasibly be obtained through self-assessment and that represent key domains related to cardiometabolic risk. This set was intended to enable a systematic evaluation of whether a limited number of simple measurements is sufficient to identify individuals at increased risk of noncommunicable diseases (NCDs) and to support scalable approaches to population-level health monitoring. To obtain such a set of biomarkers, we first reviewed the following existing health standards to determine available biomarkers and their potential relevance in the context of positive health: International consortium for health outcome measures (ICHOM) related to overall health and physical health, Health Utilities Index (HUI-mark III), International Classification of Functioning, Disability and Health (ICF), NANDA International (NANDA-I), the global organization dedicated to develop standardized nursing diagnoses and OBEsity Diverse Interventions Sharing (OBEDIS), a European research initiative to standardize the data collected in clinical trials related to obesity^14–18^. Subsequently, an expert panel comprising medical doctors (MD) and professors in the field of NCDs, physiology and epidemiology was assembled to identify relevant health domains and select representative biomarkers. Together with the experts, three health domains were identified that relate most strongly to NCD risk: metabolic health, cardiovascular health and physical fitness. These health domains are reflected by underlying physiological systems, such as glucose and lipid metabolism, fat distribution, and aerobic capacity, ie. cardio-respiratory fitness. These systems can be measured by clinical risk markers, such as circulating levels of glucose, insulin, lipid profiles and body fat percentage (Figure 1).

**Figure 1.**
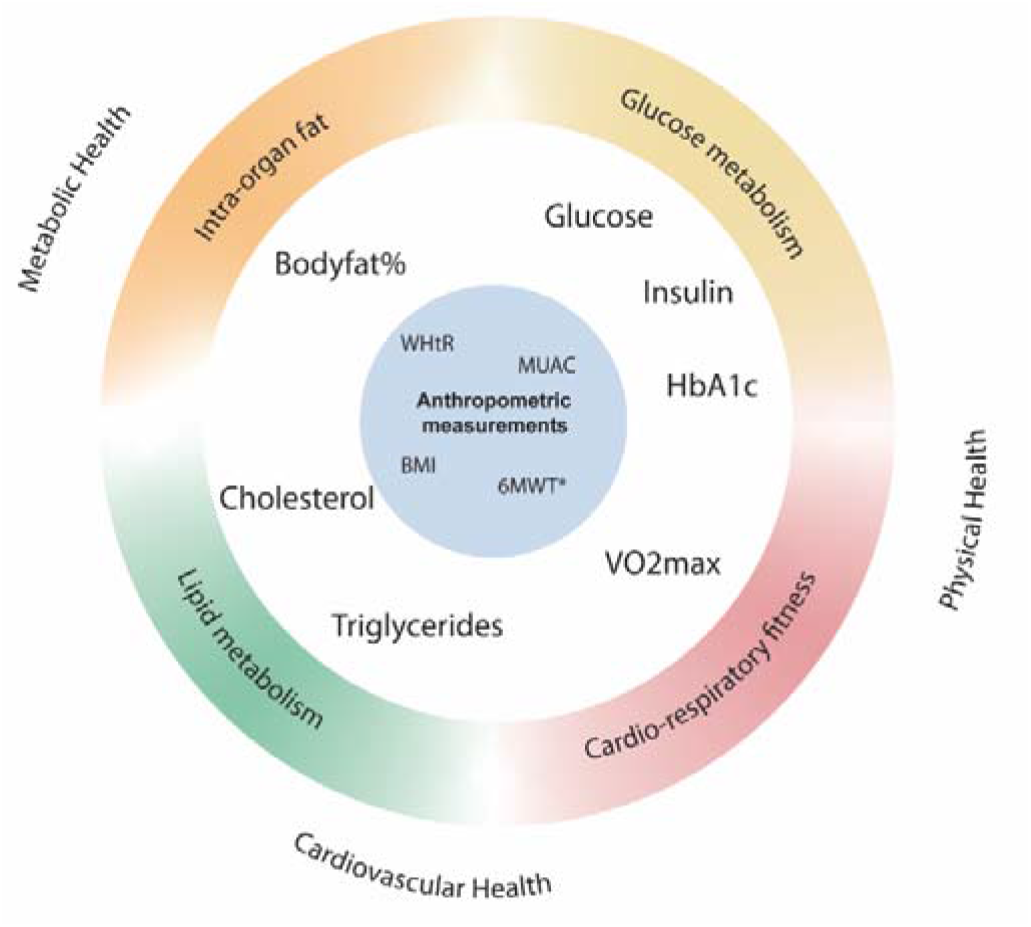
Conceptual framework of anthropometric measurements as candidate indicator and their relation to clinical risk factors and health domains. Center: Anthropometric measurements used as candidate indicators, second ring: clinical risk factors, outer ring: health markers, outside: health domains *6MWT was not available in the NHANES dataset and was therefore replaced with VO_2_max which is considered to be the golden standard for measurement of physical health.

We identified four simple, valid and objective measurements that together serve as proxy indicators for physiological health and that can be obtained at-home via self-measurement. The measurements identified were BMI as a proxy for lipid metabolism^10^, waist-to-height ratio (WHtR) as a proxy for body fat percentage^12^, mid-upper arm circumference (MUAC) as a proxy for glucose metabolism^11^, and the 6-minute walk test (6MWT) as a proxy for cardiorespiratory fitness^13^ and were selected based on health standards^14–18^. Together, they define the candidate measurement set evaluated in this study.

### 2.2. Study population (NHANES) and Patient involvement

We used data from publicly available data sets from the National Health and Nutrition Examination Survey (NHANES) from the years 1999–2000, 2001–2002, 2003–2004. In total 8275 participants that had data on biomarkers as well as outcome variables were included. Since this analysis was based on secondary data, patient involvement was not feasible. The 6MWT was not available in NHANES. However, in the cohorts 1999-2003 VO2max was measured, therefore these cohorts were selected, with VO2max as an alternative measure for 6MWT. The criteria and cut-off values used to categorize the anthropometric indicators and VO_2_max are presented in Table 1. In total, data was available from n=8275, and in a subsample with VO_2_max measurements of n=2919. Missing data were handled using outcome-specific complete-case analysis, thus resulting in varying sample sizes per outcome.

**Table 1.** Candidate indicators and respective cut-offs used for predicting clinical risk.

| **Candidate Indicator** | **Criterion** |
| --- | --- |
| BMI (kg/m<sup>2</sup>) | < 25 |
|  | (25, 30] |
|  | ≥ 30 |
| WHtR | < 0.5 |
|  | (0.5, 0.6] |
|  | ≥ 0.6 |
| MUAC (cm) | ≥ 28.4 (if age > 65) |
|  | Otherwise |
| VO<sub>2</sub>max (ml/kg/min) | < 35 (men) |
|  | < 27 (women) |
|  | Otherwise |

Cut-offs based on epidemiological data<sup>11,12,14,19</sup>

### 2.3. Outcomes

We identified endpoints available in the NHANES data that are representative of intra-organ fat, glucose and lipid metabolism. These endpoints were body fat percentage (intra-organ fat), HbA1c, glucose & insulin levels (glucose metabolism), cholesterol and triglyceride levels (lipid metabolism). Outcomes were categorized into low risk and high risk groups using cut-off values based on epidemiological data (glucose >5.6mmol/l, insulin >174pmol/l, triglycerides >1.7mmol/l, hba1c >6%, cholesterol >5mmol/l, %bodyfat > 21% for men and >31% for women)^19–22^.

### 2.4. Statistical analysis

To examine how well the defined minimal set of simple, self-measured, at-home indicators reflect underlying clinical cardiometabolic risk, we used a combination of complementary validation approaches to assess their relative and incremental contribution.

#### 2.4.1. Nested modelling approach

To identify models with minimal measurement burden, we assessed incremental performance gains across nested steps and selected models that achieved most of the improvement with fewer predictors. For each binary clinical risk outcome, we fitted multivariable logistic regression models with stepwise addition of candidate predictors. Models were nested to enable direct comparison of added predictors:

i. base model including age and sex;
ii. base+BMI;
iii. base+WHtR;
iv. base+BMI+WHtR; and
v. full model

To evaluate the additional contribution of fitness, VO2max was analyzed in participants with available VO2max measurements. Since VO2max was only measured in a subset of the population, thereby impairing comparability, we recalculated the full model in the VO2max subset (but without including VO2max) and compared it to the full model including VO2max.

Nested models were compared using likelihood ratio tests (LRTs) to test whether inclusion of additional predictors improved model fit. Discrimination was quantified using the area under the receiver operating characteristic curve (ROC-AUC). In addition, because several outcomes were imbalanced, we calculated the area under the precision–recall curve (PR-AUC; average precision) to quantify the model’s ability to prioritize true high⍰risk individuals among those assigned the highest predicted risk. As no established cut-offs exist to define clinically meaningful increases in PR-AUC, added value was interpreted in relative terms, by comparing both the magnitude of PR-AUC changes across nested models and the contribution of different measurements within the same modelling framework. In this context, predictors yielding substantially larger improvements relative to others were considered more relevant.

To additionally assess potential differences by sex, we repeated models including an interaction term for sex with candidate predictors.

#### 2.4.2. Decision trees

To account for potential non-linear relationships between predictors and cardiometabolic risk and to assess the relative importance of indicators within the predefined minimal set, we additionally applied decision tree analysis. This method takes a recursive, top-down approach, beginning with the binary outcome variable and systematically identifying the variables that best split the dataset into increasingly homogeneous subgroups. At each step, the algorithm selects the variable and threshold that most effectively separates participants based on their outcome category. Through multiple iterations, the goal is to form terminal nodes that predominantly consist of individuals belonging to a single risk category, thereby enhancing the model’s predictive accuracy. We performed exploratory sex-stratified analyses to assess whether the relative importance differed between men and women. Decision tree models were constructed using the rpart package in R^23^. To prevent overfitting, we specified a minimum split size of 10 and a minimum bucket size of 5.

## 3. Results

### 3.1. Study characteristics

Our sample consisted of 8275 individuals with a mean age of 40.5 years (Table 2). The sample included slightly more women than men (53.2%) and the biggest ethnic group was non-Hispanic whites (41.9%) followed by Mexican Americans (28.3%). With a mean BMI of 27.8 kg/m^2^ the sample was on average overweight. Furthermore, both mean WHtR (0.57) and MUAC (32.4cm) were increased in the sample. Clinical parameters were mostly in the normal range, with total cholesterol and especially body fat percentage being elevated. VO2max was measured only in a subsample of the population (n=2919). This subsample was notably younger than the overall sample (29.9 years vs. 40.5 years in the overall sample, S1).

**Table 2.** Study Characteristics of NHANES 1999-2004 participants Variable n=8275.

| Variable | n=8275 |
| --- | --- |
| Female, n(%) | 4401 (53.2%) |
| Age in years, mean±SD | 40.5±18.5 |
| Ethnicity, n(%) |  |
| Mexican American | 2338 (28.3%) |
| Other Hispanic | 463 (5.6%) |
| Non-hispanic white | 3469 (41.9%) |
| Non-hispanic black | 1700 (20.5%) |
| Other | 305 (3.7%) |
| BMI (kg/m<sup>2</sup>), mean±SD | 27.8±6.4 |
| Waist-to-Height ratio, mean±SD | 0.57±0.1 |
| Mid-upper arm circumference (cm), mean±SD | 32.4±5.0 |
| VO<sub>2</sub>max *, mean±SD | 41.1±10.6 |
| Glucose (mmol/l), mean±SD | 5.6±1.8 |
| Insulin (pmol/l), mean±SD | 82.4±79.1 |
| HbA1c, mean±SD | 5.4±1.0 |
| Triglycerides (mmol/l), mean±SD | 1.6±1.2 |
| Total cholesterol (mmol/l), mean±SD | 5.1±1.1 |
| Body fat percentage, mean±SD | 33.2±9.2 |

\*VO<sub>2</sub>max only available in a subset of n=2919

### 3.2. PR-AUC across nested models

Our nested modelling approach quantified individual contribution of each candidate indicator by comparing AUC, PR-AUC and Brier score. Across almost all models WHtR showed higher PR-AUC compared to BMI, indicating it has better discriminative performance. While accuracy (as indicated by Brier score) improved somewhat for the combination of BMI & WHtR (compared to WHtR alone), further improvements in PR-AUC were observed primarily for the outcome elevated triglyceride levels (S2). Further addition of measurements added only negligible to the classification ability.

Classification improved in the VO_2_max subsample without adding VO_2_max. Further addition of VO_2_max did not further improve classification (Figure 2).

**Figure 2.**
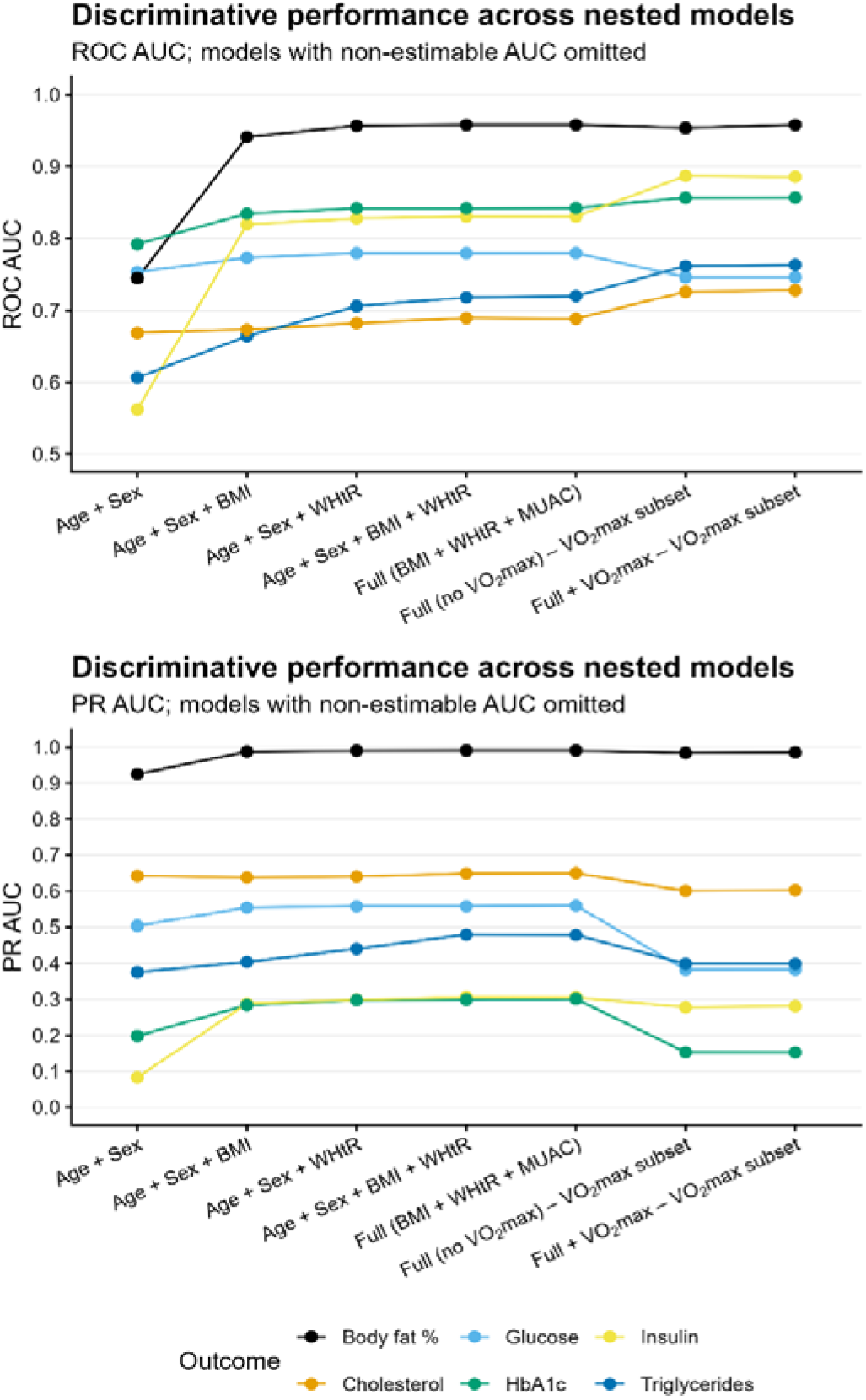
Discriminative performance of nested logistic regression models predicting elevated clinical risk factor, expressed as ROC-AUC (overall discrimination) and PR-AUC (sensitive for imbalances)

Models including an interaction term of sex with candidate predictors showed largely similar results (S3).

### 3.3. Decision trees

While regression models estimate overall associations, decision trees reveal which measurements drive classification decisions and whether risk differentiation is primarily determined by a small number of threshold⍰based splits (e.g., a single WHtR cut⍰off). Thus, we complement logistic models, which primarily assess whether adding measurements improved predictive performance, with decision trees to clarify which measurements drive classification decisions.

Decision trees could be generated for the outcomes triglycerides, total cholesterol and bodyfat (S4). Across all decision trees that could be constructed, waist-to-height ratio (WHtR) was consistently the first measurement used to separate individuals into different risk groups, indicating that initial classification was largely determined by WHtR. For body fat percentage, BMI and MUAC added modest incremental value. When compared to a model based on BMI only, improvements in sensitivity and overall accuracy were observed, mainly driven by the addition of WHtR (Table 3). Although absolute balanced accuracy differed between logistic regression (model_BMI_=87.6%; model_full incl. VO2max_=89.2%) and decision tree models, both approaches consistently identified waist-to-height ratio (WHtR) as the primary source of improvement over BMI, while additional measurements contributed only modest, outcome-specific gains.

**Table 3.** Comparison of classification performance of two decision tree models predicting body fat percentage.

| Metric | Model Full (incl. $\text{VO}_{2\text{max}}$) | Model BMI only |
| --- | --- | --- |
| Accuracy | 91.4% | 88.1% |
| Sensitivity | 76.5% | 43.9% |
| Specificity | 94.4% | 96.8% |
| Kappa | 0.694 | 0.485 |
| Balanced Accuracy | 85.4% | 70.3% |
| Detection Rate (FALSE) | 12.5% | 7.2% |
| McNemar's Test p-value | 0.0129 | < 2.2e-16 |

In summary, these findings are in line with results from logistic regression models and highlight the relevance of anthropometric measurements beyond BMI, particularly WHtR, in identifying individuals at cardiometabolic risk characterized by elevated bodyfat, triglycerides and total cholesterol.

For HbA1c, insulin and glucose no decision tree could be generated indicating that no meaningful splits could be identified that improved classification based on the available predictors and the specified model parameters.

In sex-stratified models, VO_2_max was found to be an influential variable for men, whereas MUAC seemed more important in women (S5) This points further towards sex-specific differences in how anthropometric measures related to cardiometabolic risk.

## 4. Discussion

In this study, we systematically evaluated a predefined minimal set of simple, self⍰measurable anthropometric and fitness indicators to assess how well they capture key aspects of physiological health relevant to cardiometabolic risk. Using both decision tree models and nested logistic regression allowed us to assess the relative importance of candidate measurements from complementary perspectives. Contrary to our hypothesis, the different anthropometric measures did not seem to capture different aspects of health when analyzed jointly. While decision trees identify dominant predictors, the nested regression models quantified the incremental gain in discrimination performance when adding measurements. Across most models and outcomes, waist-to-height ratio (WHtR) consistently emerged as the most influential predictor of elevated clinical risk factors, with BMI adding moderate additional value. MUAC and VO2max showed limited added value overall, although their contribution varied across outcomes and was more pronounced in sex stratified analyses. The consistent dominance of WHtR across models and outcomes is in line with previous research, which identified WHtR as an indicator for individuals with an increased cardiometabolic risk ^24,25^. The findings in this study also suggest that there seems to be an overlap of the studied anthropometric measures and the health aspects they are intended to represent. Although deriving actionable decision rules from the current analysis is limited, S6 illustrates how the observed differences in relative measurement contribution may conceptually inform future prioritization of measurements. In the future, integration of anthropometric data with digital health technologies, such as wrist-worn accelerometers, smartwatches and mobile applications could be a valuable addition by providing VO2max estimations.

VO2max played overall a negligible role in classification, except for subgroups of men. This is surprising as previous research has repeatedly shown VO2max to be a predictor of metabolic health and mortality risk^26,27^. In the study population used for this analysis, VO2max was only measured in a younger subsample of the population without pre-morbidities. As such there is likely a selection bias resulting in low variation in VO2max measurements, thereby limiting its predictive potential. We expect that replication in other datasets would likely result in VO2max playing a more important role in predicting clinical risk factors.

Decision tree analysis revealed that combining the measurements from our predefined minimal set of simple, valid, at-home indicators improved classification performance compared to using BMI alone, yielding higher sensitivity and balanced accuracy and thereby enabling detection of individuals with atypical body composition who may be missed by BMI-based screening only. This is particularly relevant in the context of Positive Health, where physiological indicators complement broader dimensions of well-being ^7^.

Exploratory decision tree analyses stratified by age and sex revealed variation in variable importance across subgroups. For example, VO2max was an important predictor primarily in men, while MUAC seemed to be more important for women. Although these differences were not the main focus of this study, they suggest that demographic factors may influence the predictive value of anthropometric measures. This is in line with multiple large-scale multi ethnic studies and meta-analysis ^28–30^. Future research including the full set of measurements could explore whether subgroup-specific models or thresholds enhance classification accuracy and clinical relevance.

The current analysis has several limitations. Due to the cross-sectional nature of the data no long-term disease risk could be taken into account. Future studies may aim to validate our models by investigating whether the proposed at-home measurements correlate with disease incidence. Secondly, in the NHANES dataset, instead of the 6MWT, VO2max was available. While this is a direct measure of aerobic fitness, it is not a measurement that can easily be done at-home. However, research has shown moderate to strong correlations between the 6MWT and VO2max, suggesting that our results would be similar with the at-home measurement^31^. Due to limited data availability and sample size, confounding factors, such as lifestyle habits, medicine use, ethnicity or socio-economic position could not be taken into account. Further, prevalence of clinical risk factors varied widely, with elevated bodyfat% being present in the majority of the sample and <10% experiencing abnormal insulin or HbA1c. Thus, in our analysis we focused on PR-AUC, a metric that accounts for class imbalances. Finally, in practice, participants would perform measurements themselves, whereas in the NHANES dataset these measurements were performed by researchers rather than the participants. Although previous literature have shown the validity of self-measured anthropometric data^32,33^, and we confirmed reliability of these measurements in a real-world efficacy study^34^ differences in measurement consistency may limit generalizability of the current findings.

In this study, we demonstrated WHtR as a robust and consistent indicator of cardiometabolic risk across several outcomes and modelling approached. Further, we showed that additional measurements only contributed marginally discriminative value in addition to WHtR. Thus, this study demonstrates, in line with previous research, that WHtR, a simple, at-home anthropometric measurement can enhance risk stratification and support the identification of individuals at risk for NCDs^35^.

Future research may extend these findings by evaluating a similar approach in longitudinal cohorts with data on disease prevalence and more complete measurement profiles to assess the contribution of different anthropometric measurements in different populations and across sexes.

## Data Availability

All data produced in the present study are available upon reasonable request to the authors

## List of Abbreviations

BMI: Body mass index
WHtR: Waist-to-height ratio
MUAC: Mid-upper arm circumference
PR-AUC: Area Under the Precision-Recall Curve
ROC-AUC: Receiver Operating Characteristic - Area Under the Curve

## 5. Declarations

### Ethics approval and consent to participate

The NHANES protocols received approval from the Ethics Review Board of the National Center for Health Statistics, with all participants providing written informed consent. Given the study’s reliance on a publicly accessible, anonymized dataset, it was exempt from further ethical review.

### Consten for publication

Not applicable

### Availability of data and materials

The datasets analysed during the current study are publicly available in the NHANES repository (https://www.cdc.gov/nchs/nhanes). The code used for analysis is available from the corresponding author on reasonable request.

### Authors’ contributions

**KS**: Writing – review & editing, Writing – original draft, Visualization, Methodology, Formal analysis. **TvdB**: Methodology, Formal analysis, Data curation, Reviewing. **MvdE**: Writing – original draft, Reviewing. **FH:** Supervision, Methodology, Reviewing. **SW:** Supervision, Methodology, Reviewing, Funding acquisition

## Acknowledgements

Not applicable

## Declaration of generative AI in scientific writing

During the preparation of this work the author(s) used Copilot in order to revise portions of the text. After using this tool/service, the author(s) reviewed and edited the content as needed and take(s) full responsibility for the content of the published article.

