## Supplement for "Minimal measurement strategies for cardiometabolic risk classification within the Positive Health framework: A cross-sectional analysis of NHANES 1999–2004 data"

| n | 2919 |
| --- | --- |
| Female, n(%) | 1404 (48.1%) |
| Age in years, mean±SD | 29.9±9.9 |
| Ethnicity, n(%) |  |
| Mexican American | 883 (30.3%) |
| Other Hispanic | 149 (5.1%) |
| Non-hispanic white | 1196 (41.0%) |
| Non-hispanic black | 594 (20.3%) |
| Other | 97 (3.3%) |
| BMI (kg/m^2^ ), mean±SD | 26.7±5.9 |
| Waist-to-Height ratio, mean±SD | 0.53±0.1 |
| Mid-upper arm circumference (cm), mean±SD | 32.1±4.8 |
| VO2max*, mean±SD | 41.1±10.6 |
| Glucose (mmol/l), mean±SD | 5.3±1.4 |
| Insulin (pmol/l), mean±SD | 73.0±59.2 |
| HbA1c, mean±SD | 5.2±0.7 |
| Triglycerides (mmol/l), mean±SD | 1.2±1.1 |
| Total cholesterol (mmol/l), mean±SD | 4.8±1.0 |
| Body fat percentage, mean±SD | 31.0±9.4 |
| Health instrument score, median [range] | 2 [0-5] |

**S1** Study characteristics in subsample with Vo2max measurement

**S2** Incremental changes in model performance across nested models

| **Outcome** | **n (full)** | **n (VO_2_max)** | **Prevalence (%)** | **ΔPR-AUC BMI vs WHtR** | **ΔPR-AUC BMIWHtR vs WHtR** | **ΔPR-AUC Full vs BMIWHtR** | **ΔPR-AUC Full (incl. VO_2_max) vs Full (excl. VO2max)** |
| --- | --- | --- | --- | --- | --- | --- | --- |
| Glucose | 3719 | 1377 | 28.3 | -0.004 | 0.000 | 0.002 | 0.000 |
| Insulin | 3658 | 1359 | 6.9 | -0.010 | 0.007 | 0.000 | 0.003 |
| HbA1c | 7573 | 2798 | 8.8 | -0.014 | 0.001 | 0.002 | 0.000 |
| Triglycerides | 3680 | 1365 | 31.0 | -0.037 | 0.039 | -0.001 | 0.000 |
| Cholesterol | 7450 | 2764 | 51.1 | -0.002 | 0.008 | 0.001 | 0.002 |
| Bodyfat% | 7317 | 2852 | 83.3 | -0.003 | 0.000 | 0.000 | 0.002 |

Incremental changes in precision–recall area under the curve (PR‑AUC). Values represent differences in performance relative to simpler reference models.
ΔPR‑AUC: positive values indicate improved discrimination

**S3** Incremental changes in model performance across nested models taking into account sex-interactions

| **Outcome** | **n (full)** | **n (VO_2_max)** | **Prevalence (%)** | **ΔPR-AUC BMI vs WHtR** | **ΔPR-AUC BMIWHtR vs WHtR** | **ΔPR-AUC Full vs BMIWHtR** | **ΔPR-AUC Full (incl. VO2max) vs Full (excl. VO2max)** |
| --- | --- | --- | --- | --- | --- | --- | --- |
| Glucose | 3719 | 1377 | 28.3 | -0.004 | 0.000 | 0.004 | 0.000 |
| Insulin | 3658 | 1359 | 6.9 | -0.002 | 0.009 | 0.000 | 0.005 |
| HbA1c | 7573 | 2798 | 8.8 | -0.009 | 0.007 | 0.001 | 0.000 |
| Triglycerides | 3680 | 1365 | 31 | -0.025 | 0.046 | 0.000 | -0.002 |
| Cholesterol | 7450 | 2764 | 51.1 | -0.003 | 0.014 | 0.003 | 0.001 |
| Bodyfat% | 7317 | 2852 | 83.3 | -0.003 | 0.001 | 0.000 | 0.001 |

Incremental changes in precision–recall area under the curve (PR‑AUC). Values represent differences in performance relative to simpler reference models.
ΔPR‑AUC: positive values indicate improved discrimination

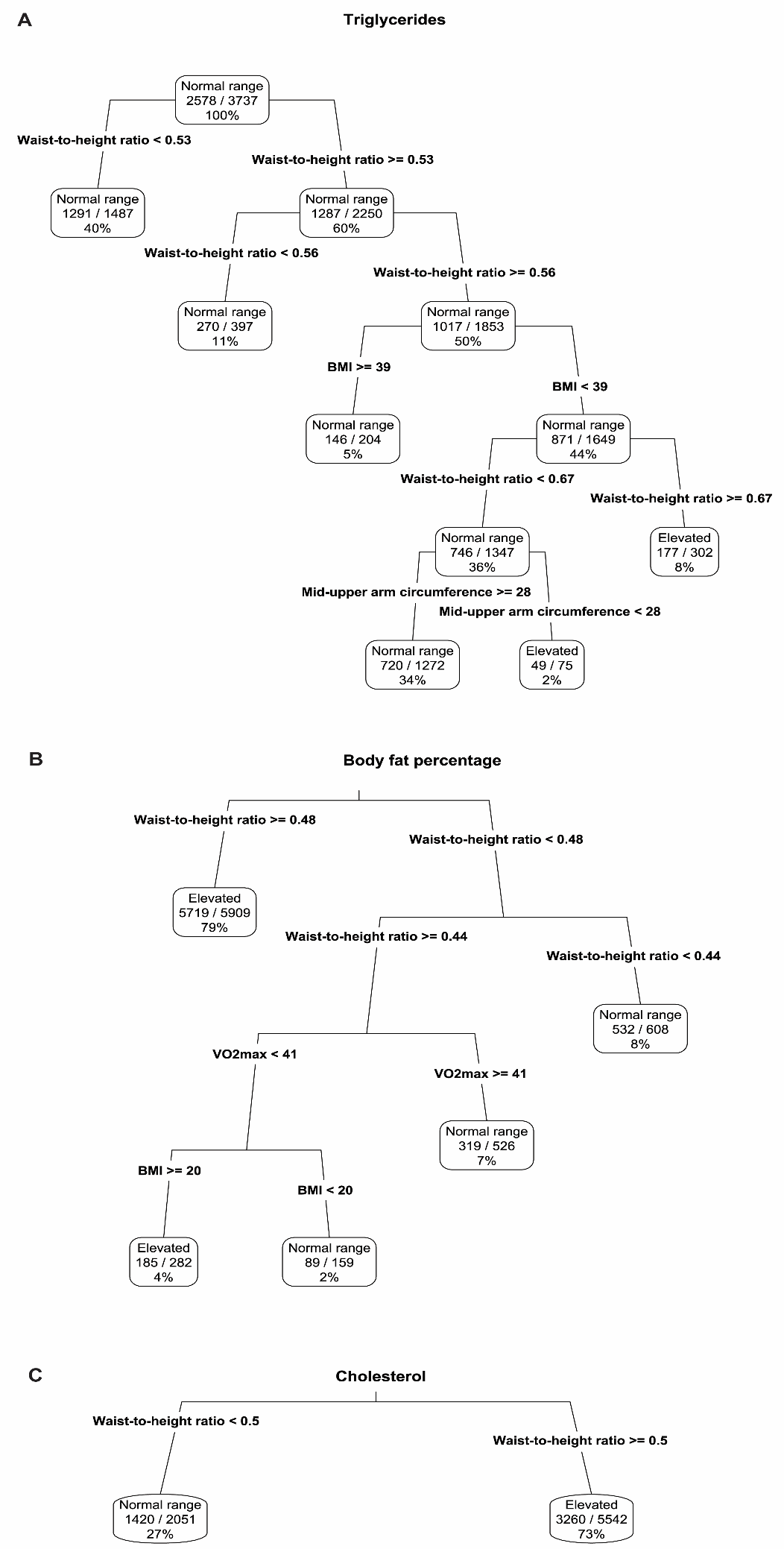

**S4** Decision trees for **A** Triglycerides, **B** Body fat percentage and **C** Cholesterol

**S5** Variable importance in sex-stratified decision trees

|  | **Triglycerides** | | **Cholesterol** | | **Body fat %** | |
| --- | --- | --- | --- | --- | --- | --- |
|  | Women | Men | Women | Men | Women | Men |
| **WHtR** | 100 | 100 | 100 | 100 | 100 | 100 |
| **BMI** | 79.8 | 84.3 | 65.9 | 60.2 | 61.2 | 51.7 |
| **MUAC** | 57.8 | 44.5 | 51.1 | 27.8 | 37.8 | 20.8 |
| **VO2max** |  | 11.0 |  |  | 1.6 | 1.3 |

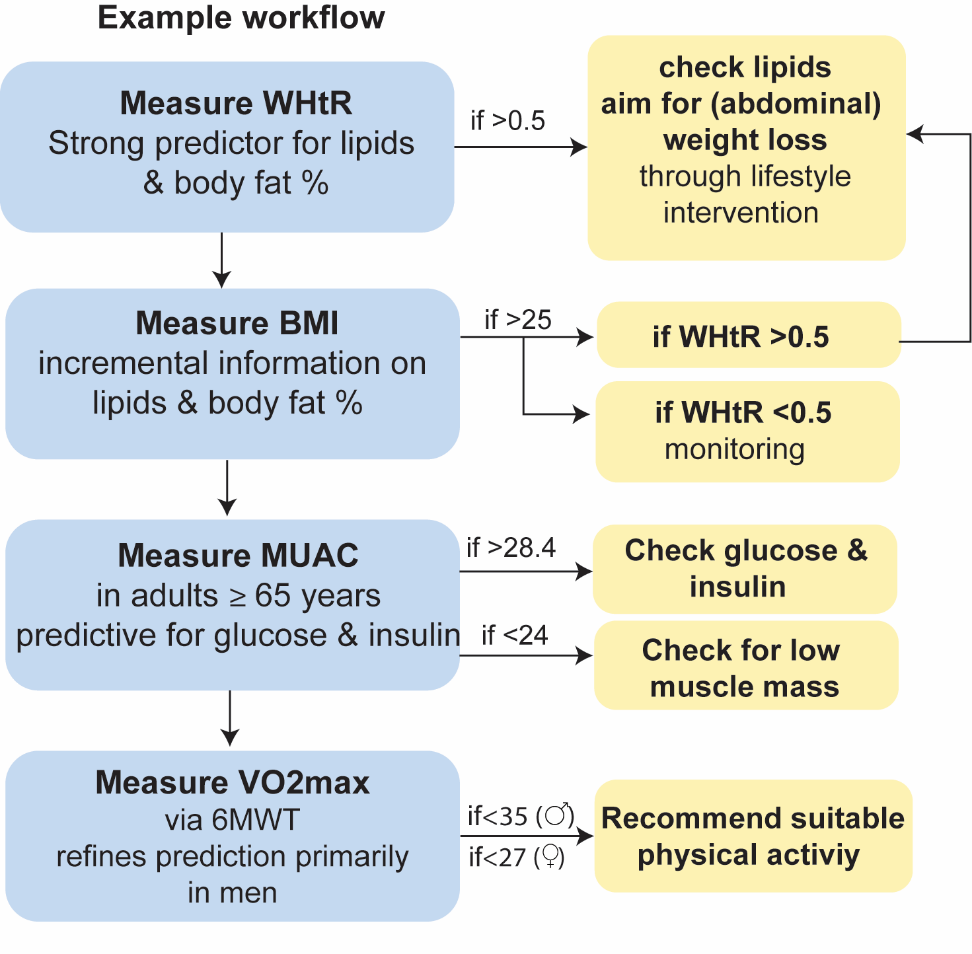

**S6 Conceptual illustration of measurement prioritization informed by relative model contributions** The ordering reflects patterns observed across nested logistic regression models and decision tree analyses, in which anthropometric measures, particularly waist‑to‑height ratio, consistently provided the largest contribution to discrimination, while additional measurements contributed more modest and context‑dependent refinements. Shown cut‑off values are based on epidemiological data. Longitudinal and external validation would be required before translation into practical application.
